# Bridging Traditional and Modern Health Systems: The Role of Digital Health in Traditional Medicine Integration in Nigeria

**DOI:** 10.1101/2025.04.29.25326680

**Authors:** Oluwafemi B. Banjo

## Abstract

This study explores the integration of traditional medicine into Nigeria’s digital health ecosystem, focusing on perspectives from patients, traditional medicine practitioners (TMPs), and medical doctors. Recognizing the enduring role of traditional medicine in Nigerian healthcare, the research assesses the feasibility and readiness for its digital relevance.

Employing a mixed-methods approach, surveys were conducted among 200 patients, 50 TMPs, and 50 medical doctors across southwestern Nigeria. Data collection emphasized demographics, awareness, usage patterns, and attitudes toward digital-traditional integration.

Findings indicate that 52% of patients are aware of digital health platforms, with 41% utilising them, primarily mobile health applications. Notably, 68% expressed willingness to engage with TMPs via digital platforms, citing convenience, easy access and cost-effectiveness as primary motivators. Among TMPs, 42% are aware of digital health platforms, yet only 19% have adopted them, hindered by limited digital literacy and infrastructural challenges. Nevertheless, 60% showed openness to digital consultations, recognizing potential benefits like broader patient reach. Medical doctors exhibited high digital health awareness (85%), with 70% actively using such tools. However, only 34% support integrating traditional medicine into modern healthcare, citing concerns over standardization, quackery and efficacy.

The study concludes that while patients and TMPs demonstrate readiness for digital integration, significant barriers persist, including infrastructural limitations and concerns from medical professionals. Addressing these challenges necessitates targeted training for TMPs, robust regulatory frameworks, and collaborative efforts to bridge traditional and modern medical practices. Such integration holds promise for enhancing healthcare accessibility and delivering holistic patient care in Nigeria.

## INTRODUCTION

In many parts of Nigeria, traditional medicine remains the primary source of healthcare for millions, particularly in rural and low-income communities. It encompasses herbal remedies, spiritual healing, bone-setting, and other indigenous practices passed down through generations. According to the World Health Organization, approximately 70 - 80% of the population in developing countries, including Nigeria, relies on traditional medicine for their basic health needs (World Health Organization, 2013). While orthodox medicine is expanding, traditional practices continue to fill the gaps left by the modern health system, especially where access to hospitals, trained personnel, and affordable treatment is limited.

Concurrently, Nigeria is experiencing rapid growth in the use of digital technologies in healthcare. Digital health innovations such as mobile health (mHealth) applications, telemedicine, electronic health records (EHRs), and artificial intelligence are improving access to care, expediting diagnoses, and strengthening health data management. These technologies offer an opportunity to better connect traditional and modern health systems, allowing for improved regulation, documentation, and integration of traditional medicine practices into the national health framework.

Despite its significance, traditional medicine in Nigeria remains largely informal, unregulated, and poorly documented. Knowledge is predominantly transmitted orally, rendering it vulnerable to loss, misuse, and misinterpretation. Furthermore, there is minimal collaboration between traditional practitioners and modern healthcare providers, leading to distrust. Digital health tools are often designed without considering traditional healthcare systems, thereby missing a valuable opportunity for synergy. Without deliberate efforts to integrate both systems, Nigeria risks losing valuable indigenous knowledge and failing to meet the health needs of many citizens.

This study provides a unique opportunity to combine the strengths of both traditional and modern healthcare systems through digital tools. Integrating traditional medicine into Nigeria’s digital health ecosystem can enhance access to culturally accepted care, support innovation in herbal and natural remedies, and promote improved health outcomes. It also facilitates the preservation of indigenous knowledge while building trust, transparency, and collaboration within the health sector.

The primary objective of this research is to explore how digital health can support the integration of traditional medicine into Nigeria’s formal healthcare system. Specifically, it aims to:

- Examine the current state of traditional medicine and digital health in Nigeria.
- Identify opportunities for synergy and integration.
- Highlight challenges and barriers to integration.
- Recommend policy and practical strategies for implementation.

## LITERATURE REVIEW

### Global and Local Perspectives on Traditional Medicine

Traditional medicine continues to play a crucial role in global healthcare, especially in low□ and middle□income countries like Nigeria. In China and India, Traditional Chinese Medicine (TCM) and Ayurveda have been successfully integrated into national healthcare frameworks through legislation, research centers, and technological platforms (World Health Organization, 2013; Senah & Kpobi, 2019). In Ghana, for example, the establishment of institutional structures such as the Centre for Plant Medicine Research (CPMR) and Traditional Medicine Practice Council (TMPC), as well as its integration into 55 government hospitals, exemplify this successful model (MoH, 2021; Esan et al., 2018).

In Nigeria, traditional medicine, including herbal remedies, spiritual healing, and bone-setting is a primary healthcare option for over 70% of the population, particularly in rural areas where access to biomedicine is limited (Adefolaju, 2014; Ibrahim et al., 2023). However, its development is hindered by the lack of formal documentation, regulation, and integration into the health system. The informal and oral nature of knowledge transmission raises concerns about safety, efficacy, and sustainability (Ozioma & Chinwe, 2019).

Although the Nigerian government has instituted a Traditional, Complementary, and Alternative Medicine (TCAM) department within the Federal Ministry of Health and revised policy frameworks, implementation remains fragmented (Adigwe, 2023; Akangbe, 2024). Scholars emphasize that integration efforts are frequently ad hoc and rely on the goodwill of supportive individuals rather than systematic policy enforcement (Ugwu, 2024).

Over the past decade, Nigeria’s digital health ecosystem has expanded rapidly. Key technologies include mobile health (mHealth), telemedicine, electronic health records (EHRs), AI diagnostics, and wearable devices (Ofori-Atta et al., 2022). These tools have enhanced hospital efficiency, remote healthcare access, disease surveillance, and public health communication—especially during emergency situations (NIPRD, 2022).

Despite the value of these developments, most digital health solutions in Nigeria are designed for biomedical contexts and fail to accommodate or incorporate traditional medical practices (Akangbe, 2024). This represents a significant gap given the prevalence and cultural embeddedness of traditional healthcare.

Emerging initiatives across Africa show promise in digitizing and integrating traditional medicine:

Ghana: The CPMR has taken active steps to digitize medicinal plant records and foster clinical research into herbal medicine (MoH, 2021; Esan et al., 2018).

Senegal: Pilot mobile initiatives document traditional practices and support hybrid consultation models (Ae-Ngibise et al., 2024).

Nigeria: The HERBINFO initiative by NNMDA aims to create a digital repository of local medicinal plants, aligning with WHO’s Traditional Medicine Strategy (Adefolaju, 2014; WHO, 2013).

Bioinformatics in Akwa Ibom: Researchers Imeh Umoren et□al. (2023) used machine learning to combine patient responses to both traditional and modern therapies, indicating a move toward evidence-based integration (Umoren et al., 2023).

Despite these initiatives, integration remains fragmented, characterized by underfunding, insufficient digital infrastructure, and a lack of intersectoral coordination (Adigwe, 2023; Akangbe, 2024).

### Key Barriers & Enablers to Integration

1. Regulatory Challenges: Poor funding and weak regulatory frameworks limit traditional medicine’s growth and acceptance. Existing initiatives (e.g., NIPRD, NAFDAC, TCAM) are promising but need systemic support (Adigwe, 2023).

2. Trust & Training: Studies across West Africa identify trust deficits and the need for cross-training between traditional and conventional practitioners as crucial for successful integration (Ae-Ngibise et al., 2024; Ofori-Atta et al., 2022).

3. Digitization Needs: Digitization can preserve indigenous knowledge, enable standardization, and facilitate integration into health information systems but requires robust infrastructure and stakeholder engagement (Akangbe, 2024; Umoren et al., 2023).

4. Utilization in Emergencies: During disease outbreaks (e.g., COVID□19, cholera), many African communities turned to traditional remedies, highlighting their potential in supportive disease response systems (Ugwu, 2024).

Though traditional medicine enjoys widespread use and cultural acceptance, it remains marginalized in Nigeria’s digital health platforms. There is a clear demand for coordinated, evidence-based, and digital-first integration frameworks, informed by global best practices (China, India) and reinforced by country-specific data and stakeholder collaboration.

## METHODOLOGY

This study adopts a quantitative research design to assess the perspectives and attitudes of patients, traditional medicine practitioners, and medical doctors toward the integration of traditional medicine with digital health platforms in Nigeria. The primary goal is to quantify the levels of awareness, usage, and acceptance of digital health tools concerning traditional medicine among various healthcare stakeholders.

### Population and Sample

The study population comprises three key groups involved in healthcare delivery: patients, traditional medicine practitioners, and medical doctors. Data were collected from a total of 300 individuals:

200 patients/individuals: These are individuals who have utilized either traditional or modern healthcare services. They represent the end-users of health services.

50 traditional medicine practitioners: This group includes herb sellers, herbalists, spiritual healers, and other traditional healthcare providers operating at the community level.

50 medical doctors: These healthcare professionals work across various levels of healthcare, including primary, secondary, and tertiary institutions, as well as private hospitals. Their views help to understand the medical community’s perception of integrating traditional medicine with modern digital health tools.

Different survey questions were administered to each category to obtain detailed information.

Data Collection Instruments

Data were collected using a structured printed questionnaire designed to assess the following key areas:

Awareness: The level of awareness regarding digital health platforms and their potential integration with traditional medicine.

Usage: The frequency of use of digital tools (e.g., mobile health apps, telemedicine services) for healthcare purposes.

Acceptance: The willingness to adopt digital health solutions to access or promote traditional medicine services.

Challenges: Identification of barriers to integration, including cultural, technical, and policy-related obstacles.

Data Collection Procedure

Data collection was conducted over a one-month period.

Questionnaires were distributed to individuals in comfortable settings, including public areas, worship centres and retail pharmacies. Traditional medicine practitioners were approached at their practice sites and during general meetings, while medical doctors were surveyed at their respective hospitals.

### Data Analysis

Most important of the collected data were analyzed and compiled using pie charts and figures to facilitate comprehensive interpretation.

### Ethical Considerations

Informed consent was obtained from all participants, ensuring they understood the study’s objectives and their right to privacy. All responses were kept confidential and anonymized to protect participants’ identities. This approach aligns with ethical standards and complies with Nigeria’s Data Protection Act (NDPA) 2023.

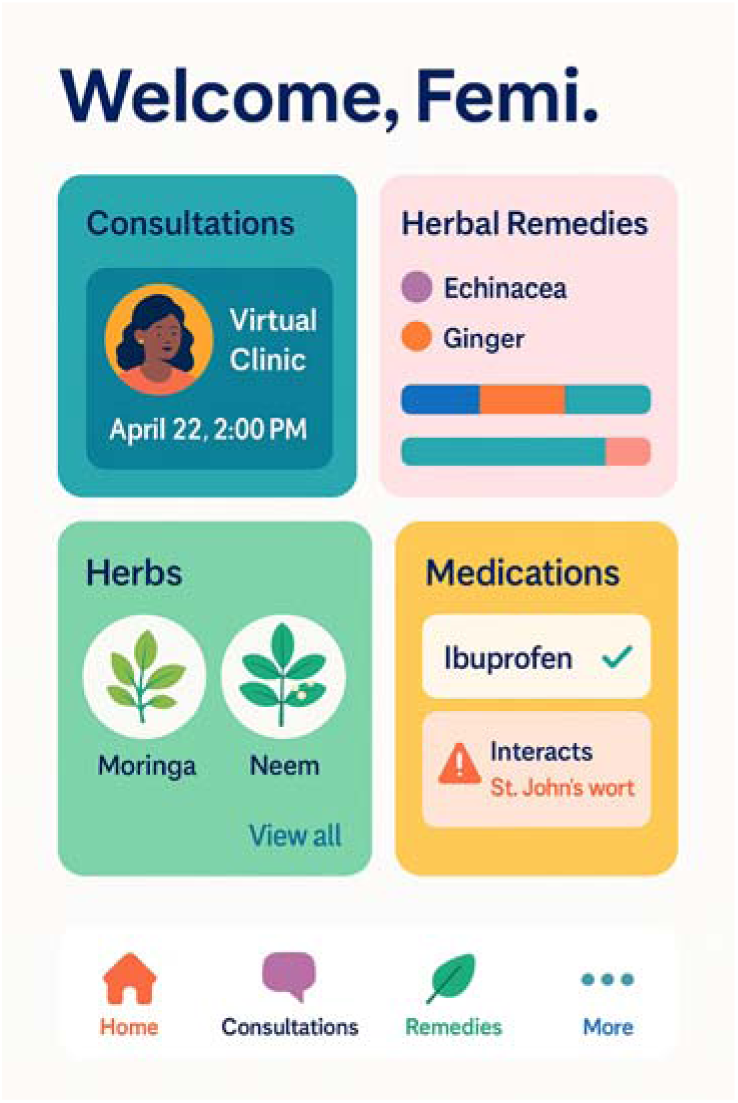
This is an AI generated Image to show what the dashboard of a traditional digital platform should look like.

## FINDINGS AND RESULTS

This section presents the analyzed responses from 200 patients/users, 50 traditional medicine practitioners (TMPs), and 50 medical doctors across various healthcare levels.

### Patients/Users’ Responses

#### Demographics

- The majority of respondents (45%) were aged 31–40, with females representing 58% of the sample.
- Educational levels varied, with 60% having tertiary education and 30% secondary education.
- 40% accessed healthcare via private hospitals, 25% via traditional medicine practitioners, and the rest through public health facilities.

#### Awareness and Usage of Digital Health

- 52% of patients were aware of digital health tools such as mobile apps and telemedicine.
- 41% had used at least one digital health tool, with mobile health apps being the most common (60%).

#### Usage of Traditional Medicine

- 62% reported occasional use of traditional medicine, while 20% used it regularly.
- Most accessed these services through direct visits to traditional healers (70%).

#### Attitudes Toward Integration

- 68% were willing to use a digital platform to consult with traditional medicine practitioners.
- Key motivations included:

- Convenience and accessibility (40%)
- Cost-effectiveness (25%)
- Trust in digital platforms (15%)
- Concerns raised included:

- Privacy and security (30%)
- Lack of trust in traditional remedies (25%)
- Low awareness about digital platforms (20%)

### Traditional Medicine Practitioners’ Responses

#### Demographics

- 50% had practiced for over 10 years; herbal medicine was the dominant speciality (60%).
- 48% had only primary/secondary education, while 21% had tertiary-level education.

#### Awareness and Usage of Digital Health

- 42% were aware of digital health platforms, but only 19% had used them.
- Tools used included:

- Social media platforms (40%)
- Telemedicine (10%)
- Health websites/apps (5%)

#### Integration Perspectives

- 60% expressed willingness to use digital platforms for consultations.
- Main challenges included:

- Limited digital skills (40%)
- Internet/device access issues (30%)
- Cultural resistance (15%)
- Perceived benefits:

- Broader patient reach (50%)
- Better patient management (20%)
- Easier collaboration with doctors (15%)

#### Attitudes Toward Adoption

- 40% were somewhat comfortable with digital health tools, and 20% were very comfortable.
- Key motivators for adoption:

- Government or NGO support (30%)
- Training and technical support (25%)
- Patient demand (20%)

### Medical Doctors’ Responses

Demographics

- 40% of respondents were general practitioners; 30% worked in private hospitals, and 40% in tertiary facilities.
- 60% had practiced for 5–10 years.

Digital Health Awareness and Usage

- 85% were familiar with digital health tools, with 70% actively using them in practice.
- Telemedicine and mobile applications were the most commonly utilized tools.

Perception of Traditional Medicine

- 34% believed that traditional medicine could be integrated into modern healthcare.
- 53% occasionally encountered patients who used traditional remedies alongside conventional care.

Integration Readiness

- 54% supported the integration of traditional and digital healthcare systems; 20% were uncertain.
- Key challenges identified included:

- Lack of standardization and regulation (40%)
- Concerns regarding efficacy and safety (30%)
- Knowledge gaps among healthcare professionals (15%)
- Perceived benefits encompassed:

- Enhanced access in rural areas (35%)
- Improved collaboration with traditional medicine practitioners (25%)
- Provision of holistic patient care (20%)

Key Takeaways

1. Patients Are Receptive to Digital Integration of Traditional Medicine

The data indicates that 68% of patients are willing to consult traditional medicine practitioners via digital platforms. While awareness of digital health tools is relatively high (52%), actual usage remains limited, highlighting a gap between exposure and active engagement. This proves the need for public sensitization campaigns, particularly targeting older adults and rural populations.

2. Traditional Medicine Practitioners Exhibit Interest but Face Digital Barriers

60% of traditional medicine practitioners (TMPs) are open to offering their services digitally; however, only 19% currently utilize digital tools, predominantly through informal channels such as WhatsApp or Facebook. Barriers include limited digital literacy, affordability of devices, and apprehension regarding regulatory implications.

TMPs are not resistant to innovation; rather, they require tailored training, incentives, and digital infrastructure to effectively participate in integrated healthcare systems.

3. Medical Doctors Cautiously Support Integration Under Specific Conditions

While not entirely opposed to traditional medicine, 34% of medical doctors would support its integration into modern healthcare, provided there are established safety regulations and scientific validation of remedies. Primary concerns include potential herb-drug interactions, the absence of clinical guidelines, and the risk of patient misinformation.

Medical professionals acknowledge the role of traditional medicine but advocate for its proper evaluation and integration to ensure patient safety.

4. Digital Integration of Traditional Practices Supports Universal Health Coverage (UHC)

The continued reliance on traditional medicine by nearly half of the Nigerian population positions it as a critical component of UHC strategies. Digitizing access to these services can address health equity gaps, particularly in under-resourced rural areas.

Integrating traditional practices with the formal health sector through digital means is a strategic approach toward achieving inclusive, people-centred UHC in Nigeria.

## DISCUSSION

This study confirms the readiness, perceptions, and usage patterns of digital health tools among patients, traditional medicine practitioners (TMPs), and medical doctors in Nigeria, with the aim of assessing the feasibility of integrating traditional and modern health systems through digital platforms. The findings reveal key trends, challenges, and opportunities with implications for policy, innovation, and future research.

### Summary of Key Findings

The data indicate that a significant proportion of patients (52%) are aware of digital health tools, with 41% having used at least one. There is substantial openness to integrating traditional medicine with digital tools, as 68% expressed willingness to consult traditional practitioners digitally. TMPs demonstrated moderate awareness (42%) and low current usage (19%) of digital tools but showed strong willingness (60%) to adopt them for broader reach and improved patient management. Medical doctors exhibited the highest digital health awareness (85%) and usage (70%), though only 34% supported the integration of traditional medicine into modern care. Nevertheless, 54% expressed support for such integration through digital means.

### Interpretation of Findings

Patients’ readiness to engage with traditional medicine via digital platforms suggests growing trust in technology-driven health solutions, particularly due to their convenience, affordability, and accessibility. However, concerns regarding data privacy, trust in traditional remedies, and limited awareness of digital platforms underscore the need for targeted education, regulation, and infrastructure development.

Among TMPs, low digital engagement is likely influenced by limited formal education and digital literacy. Nonetheless, the perceived benefits of broader patient access indicate that, with appropriate training and institutional support, adoption could increase significantly. The willingness of TMPs to collaborate digitally with medical professionals reflects a progressive attitude towards integrative healthcare.

Medical doctors, while well-versed in digital health applications, remain cautious about the integration of traditional medicine. Their concerns, centred on regulation, standardization, and clinical efficacy, highlight the necessity of scientific validation of traditional practices. However, their recognition of potential benefits, including improved rural access and holistic care, indicates that structured frameworks could foster greater acceptance.

### Comparison with Existing Literature

These findings align with global trends outlined by the World Health Organization (WHO, 2022), which advocates for the role of digital health in unifying traditional and modern care systems, particularly in low and middle-income countries. Comparable studies in East and Southern Africa (Ongolo-Zogo et al., 2020) have similarly identified that, while skepticism persists, digital collaboration models can improve patient-centred care and health outcomes in underserved populations.

### Implications for Health System Integration

The results proves the potential of digital health as a transformative bridge between traditional and medical healthcare systems. For policymakers and investors, this study provides evidence to support the development of inclusive digital platforms that are multilingual, culturally sensitive, and accessible across varying literacy levels. Integration models should incorporate regulated entry pathways for TMPs, including certification, ethical guidelines, and collaborative referral systems involving medical professionals.

For digital health innovators, the findings highlight a market demand for user-friendly technologies tailored to the needs of TMPs and their clientele, especially in rural settings where traditional medicine remains predominant. For healthcare professionals and institutions, capacity-building programs and interdisciplinary collaboration can foster trust and mutual respect between the health systems.

### Limitations

This study has several limitations. While the sample size (300 respondents) is significant, the geographic scope was limited to selected regions, which may not comprehensively reflect Nigeria’s diverse population. Finally, the study assessed perceptions and readiness rather than the actual impact of integration, which would require a longitudinal research design.

## CONCLUSION

This study has examined the evolving progress of healthcare in Nigeria through the lens of digital health integration with traditional medicine. By triangulating insights from patients/users, traditional medicine practitioners (TMPs), and medical doctors, the research assessed their levels of awareness, usage patterns, perceptions, and readiness for a digitally integrated healthcare system.

The findings indicate a growing awareness of digital health tools among all stakeholders, albeit with disparities in adoption rates, particularly among TMPs. While patients/users demonstrated a strong willingness to utilize digital platforms to access traditional care, motivated by convenience and affordability, TMPs faced challenges related to digital literacy and infrastructural limitations. Conversely, medical doctors expressed cautious optimism, supporting integration contingent upon standardization, regulation, and evidence-based validation of traditional remedies.

This divergence in readiness confirms a broader issue: the integration of indigenous and modern healthcare systems cannot rely solely on technological solutions. It must go in hand with policy reforms, capacity building, trust-building across healthcare disciplines, and cultural sensitivity.

The study also raises a fundamental question; can technology bridge the historic divide between two parallel yet potent health systems? The data suggest it can, but only through deliberate efforts by government, academia, tech innovators, and healthcare stakeholders.

In a country where over 70% of the population continues to rely on traditional medicine, failing to integrate it into the formal healthcare system, especially through digital means, risks marginalizing millions. Digital platforms present a transformative opportunity to democratize access, standardize care, and foster collaboration between knowledge systems that have long operated in isolation.

Therefore, integrating traditional medicine into digital health platforms in Nigeria will represent a socio-cultural revolution, one that demands a shared vision, mutual respect, and the strategic application of digital innovation to construct a more inclusive healthcare system for the future.

## RECOMMENDATIONS

Based on the findings and discussion, the following recommendations are proposed:

### 1. Policy and Regulatory Frameworks

The Federal Ministry of Health, in collaboration with the National Information Technology Development Agency (NITDA) and the National Agency for Food and Drug Administration and Control (NAFDAC), should develop a comprehensive policy framework for the digital integration of traditional medicine. This framework should encompass standards for safety, documentation, and practitioner accreditation.

### 2. Capacity Building for TMPs

Implement nationwide training programs to enhance digital literacy among traditional medicine practitioners, covering areas such as mobile health application usage, electronic record-keeping, patient data management, and remote consultation techniques.

### 3. Investment in Digital Infrastructure

Enhance internet accessibility and affordability in underserved regions to facilitate digital consultations and data sharing between TMPs and formal healthcare institutions.

### 4. Collaborative Research and Innovation

Encourage joint research initiatives between medical scientists/NAFDAC and TMPs to validate traditional remedies and incorporate verified treatments into digital formularies and healthcare platforms.

### 5. Public Awareness Campaigns

Design culturally sensitive campaigns to educate the public on the benefits and safety of integrating traditional medicine with digital health solutions, addressing prevalent myths, fears, and misconceptions.

### 6. Pilot Integration Platforms

Develop and evaluate pilot digital health platforms that connect TMPs, medical doctors, and patients. These pilots can serve as models for learning and refinement before nationwide implementation.

## Supporting information

Survey chat. Supplemental

## Data Availability

All data produced in the present work are contained in the manuscript

## ETHICAL CONSIDERATIONS

This study adhered to ethical standards of voluntary participation, confidentiality, and informed consent. All participants were briefed on the purpose of the study and assured that their responses would be anonymized and used solely for research purposes.

No sensitive data was collected beyond the scope of the survey instruments, and participants had the right to withdraw at any point.

