## Supplementary material for "Bridging Traditional and Modern Health Systems: The Role of Digital Health in Traditional Medicine Integration in Nigeria": Survey chat. Supplemental

### Charts: Findings from Survey

#### Age Distribution (Patients)

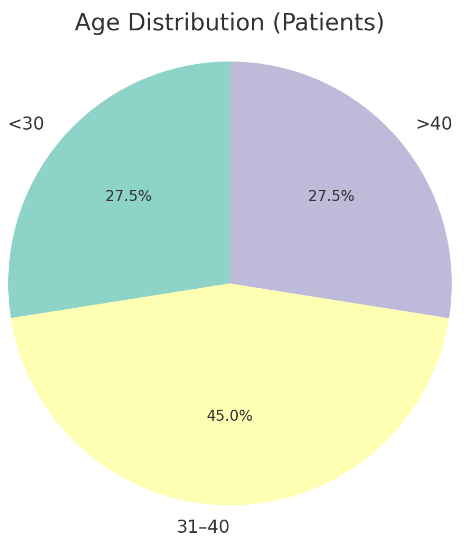


#### Gender Distribution (Patients)

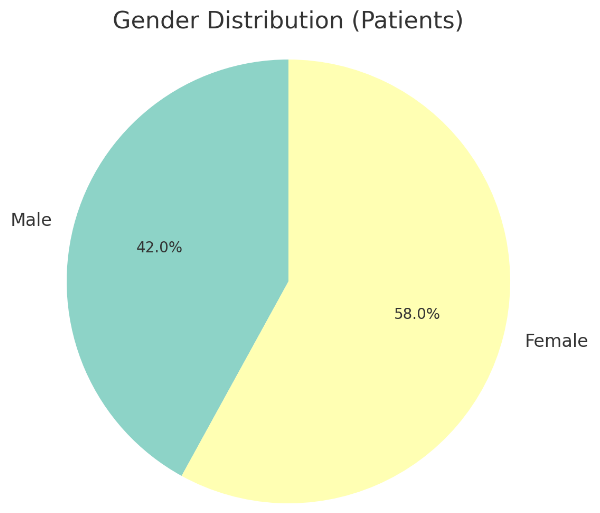

#### Education Level (Patients)

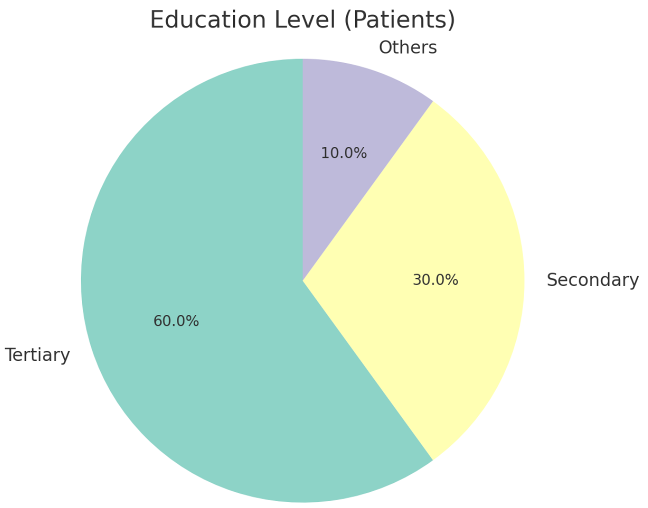


#### Healthcare Access Points (Patients)

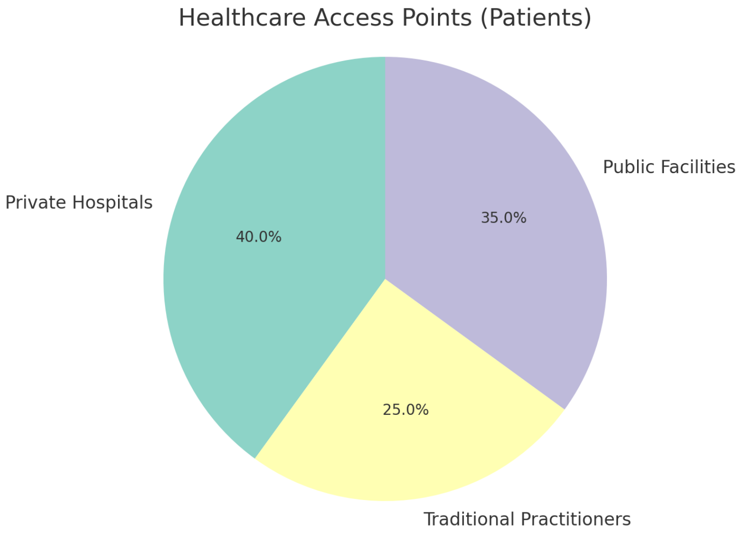

#### Awareness of Digital Health (Patients)

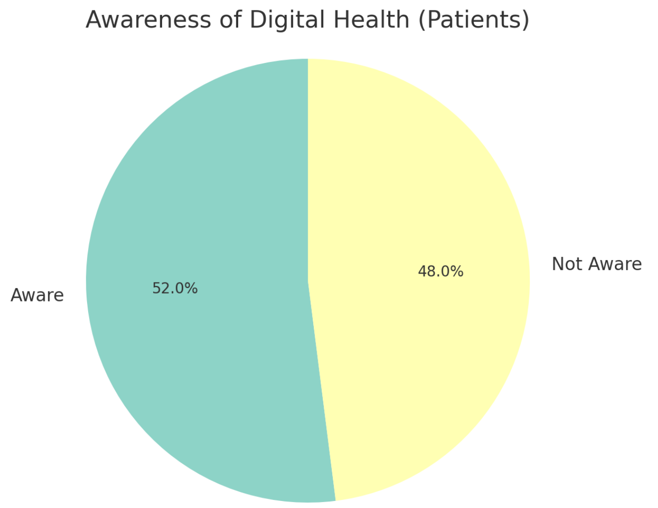


#### Usage of Digital Health Tools (Patients)

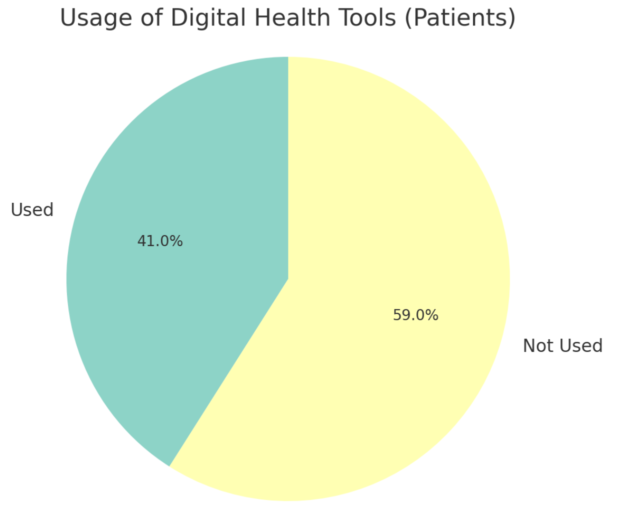

#### Type of Digital Tools Used (Patients)

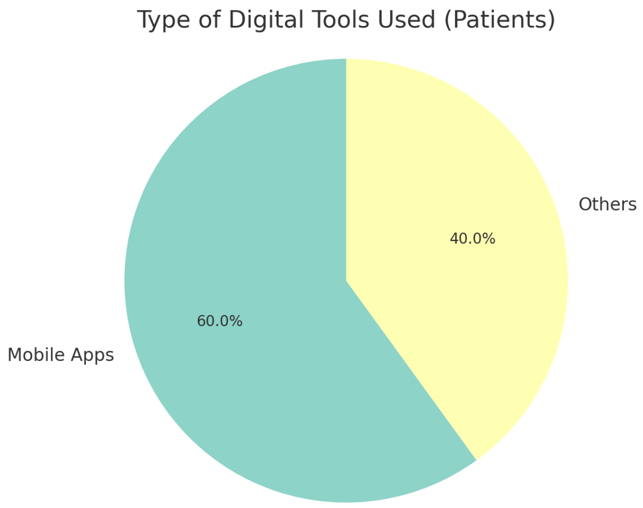


#### Frequency of Traditional Medicine Use (Patients)

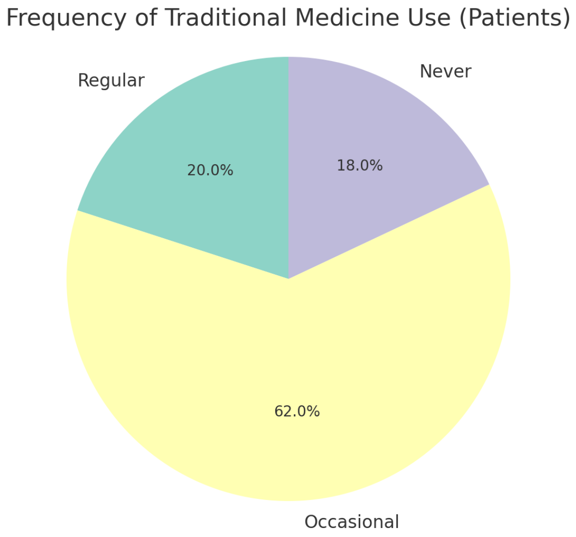

#### Mode of Accessing Traditional Medicine (Patients)

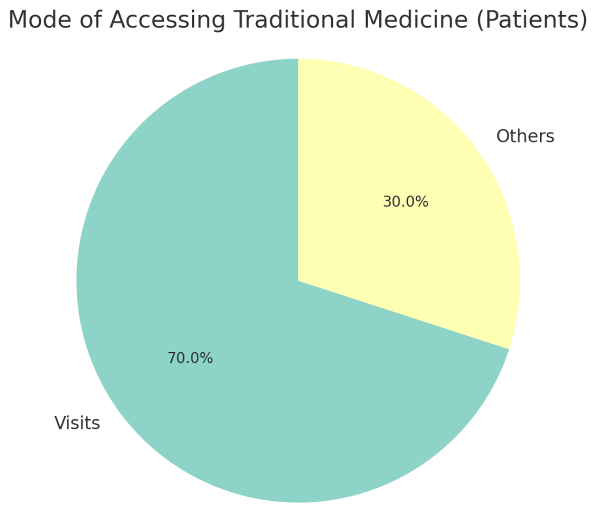


#### Willingness to Use Digital Platforms (Patients)

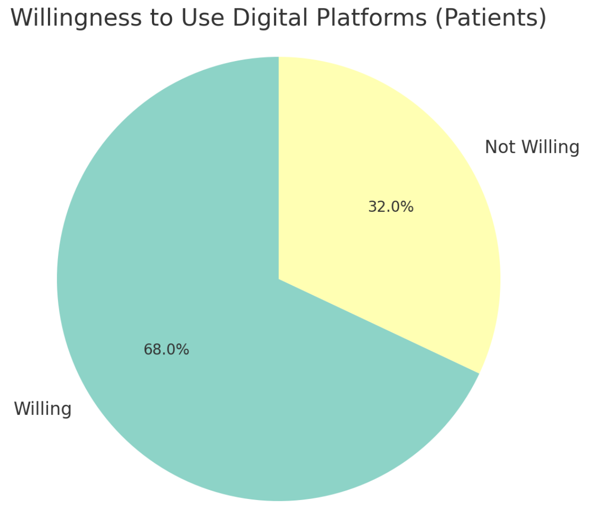

#### Motivations for Using Digital Platforms (Patients)

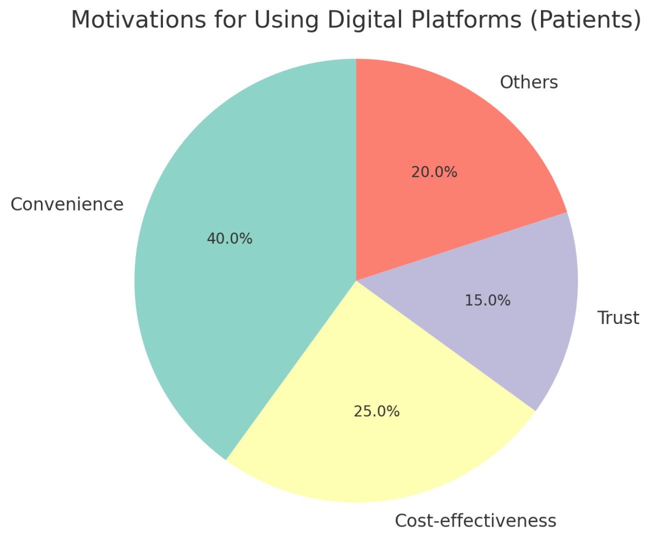


#### Concerns Regarding Digital-Traditional Integration (Patients)

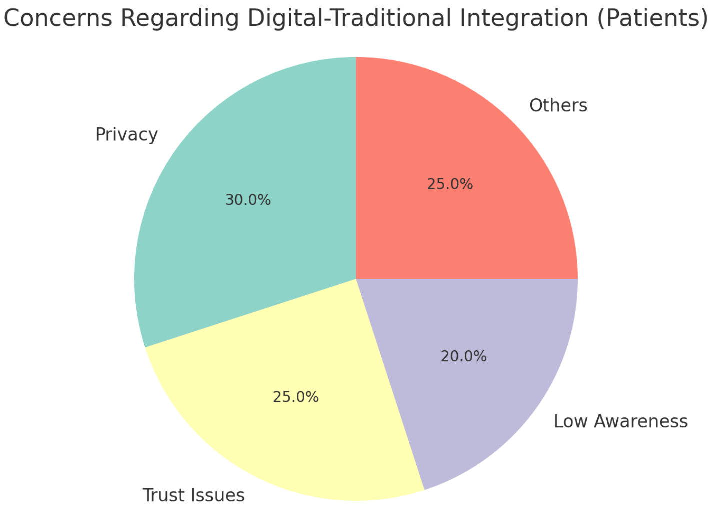

#### Years of Experience (TMPs)

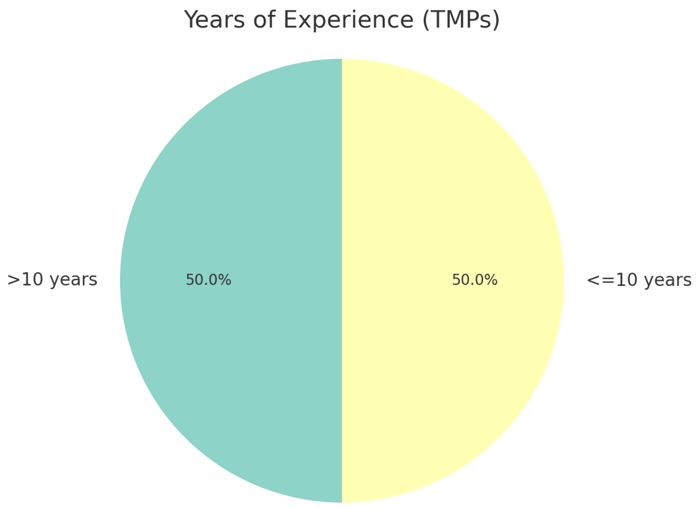


#### Specialty Area (TMPs)

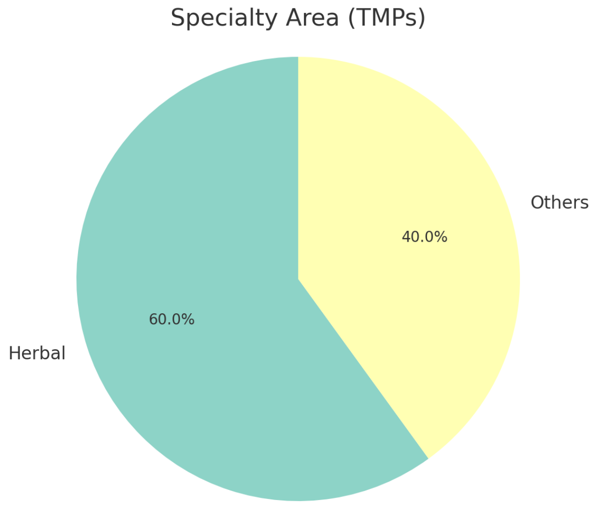

#### Education Level (TMPs)

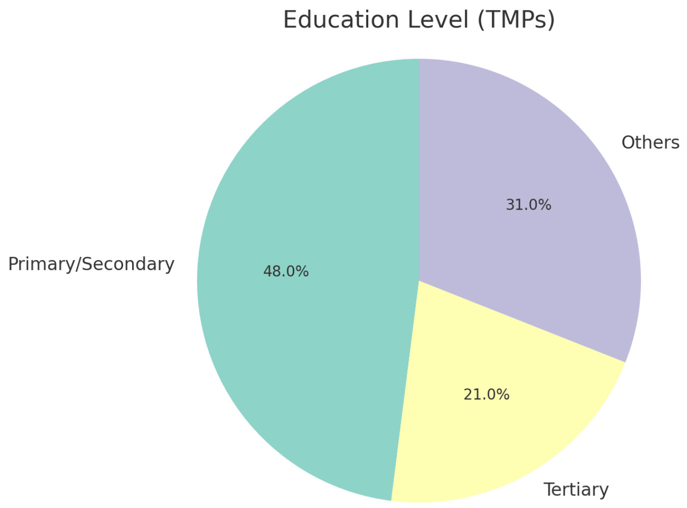


#### Awareness of Digital Health (TMPs)

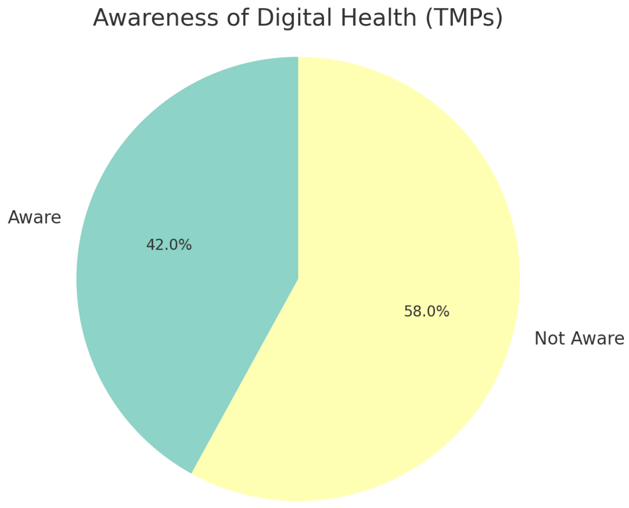

#### Usage of Digital Tools (TMPs)

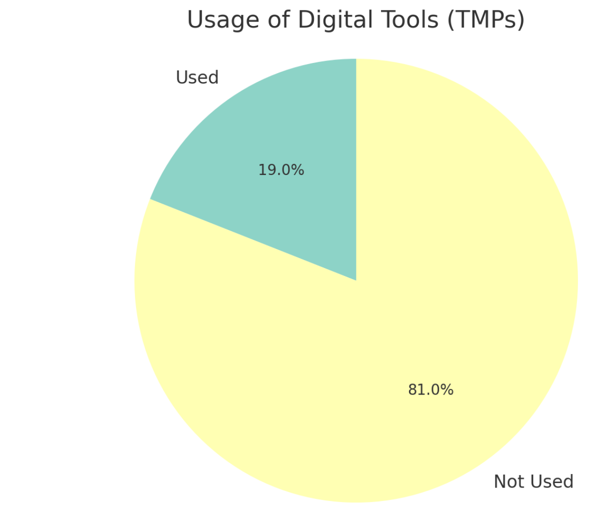


#### Type of Digital Tools Used (TMPs)

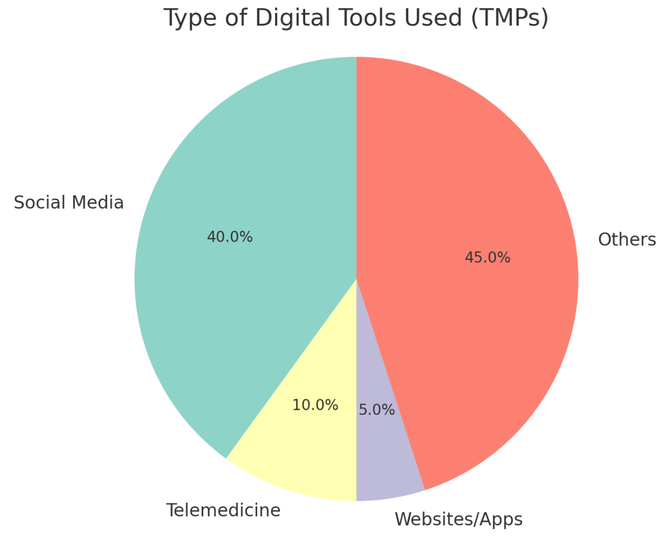

#### Willingness to Integrate Digital Platforms (TMPs)

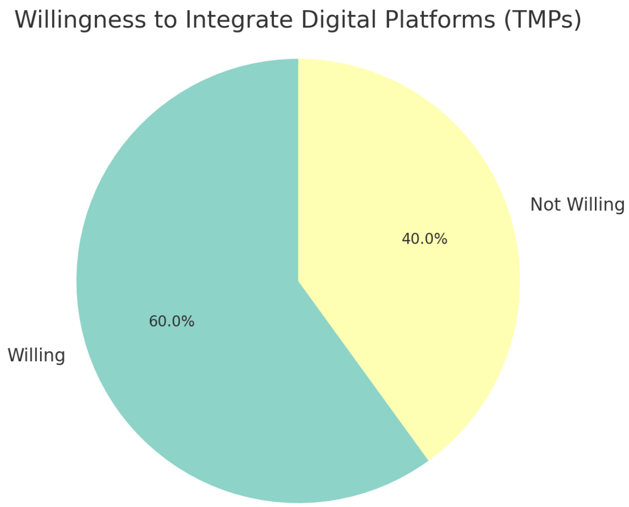


#### Challenges to Digital Use (TMPs)

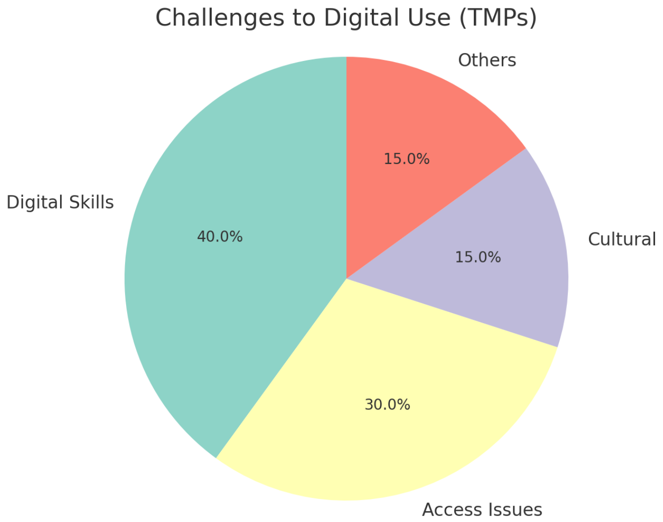

#### Perceived Benefits of Digital Integration (TMPs)

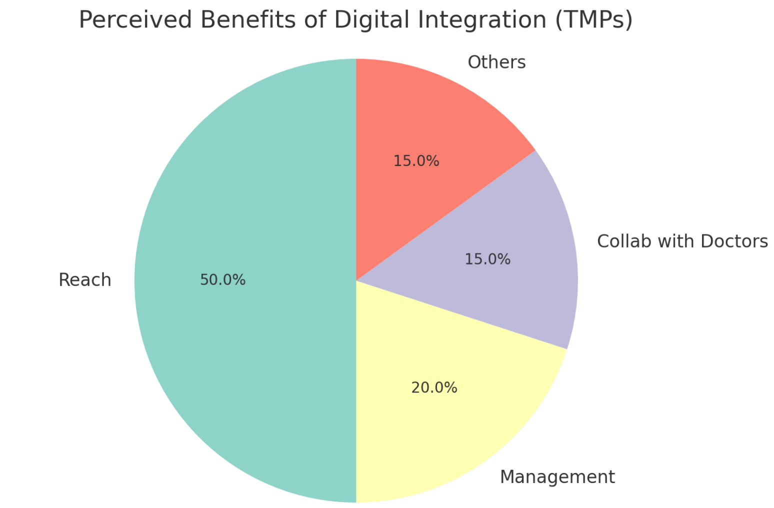


#### Comfort Level with Digital Health Tools (TMPs)

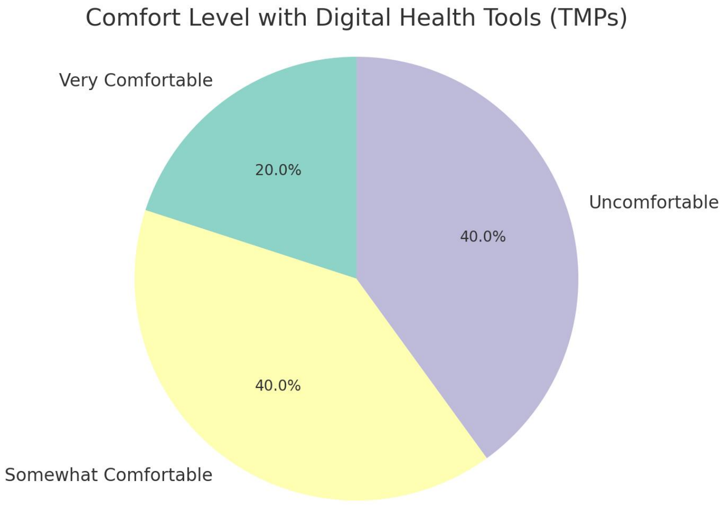

#### Motivators for Adoption (TMPs)

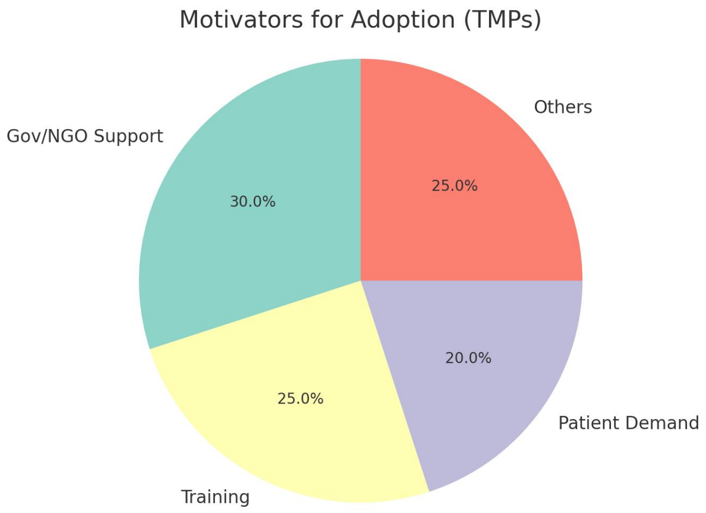


#### Role of Respondents (Doctors)

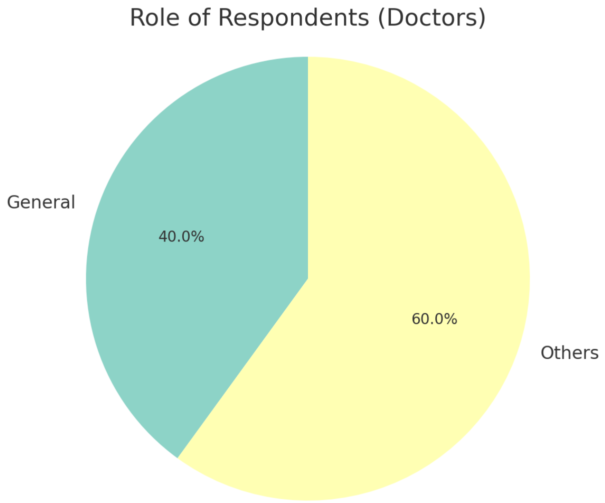

#### Workplace Setting (Doctors)

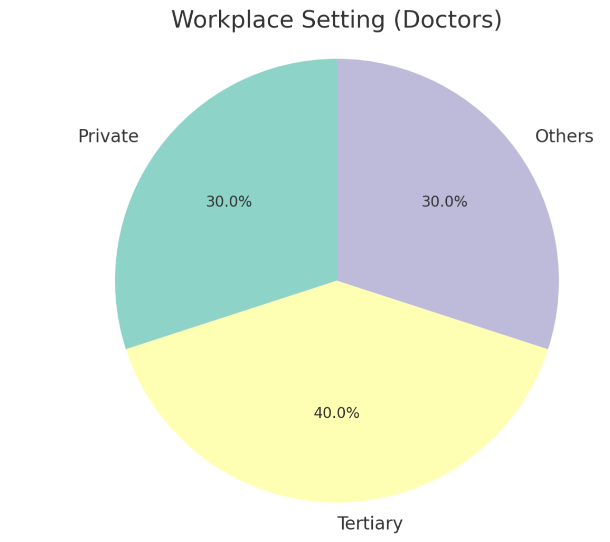


#### Years of Practice (Doctors)

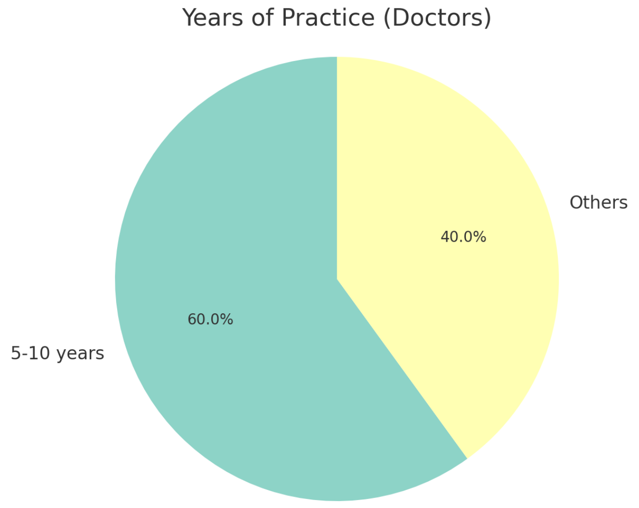

#### Familiarity with Digital Health (Doctors)

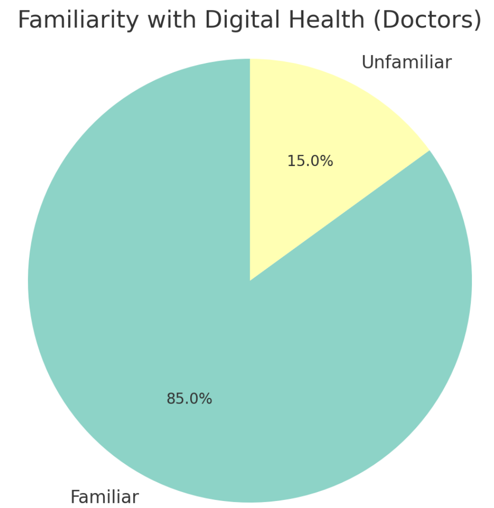


#### Usage of Digital Tools (Doctors)

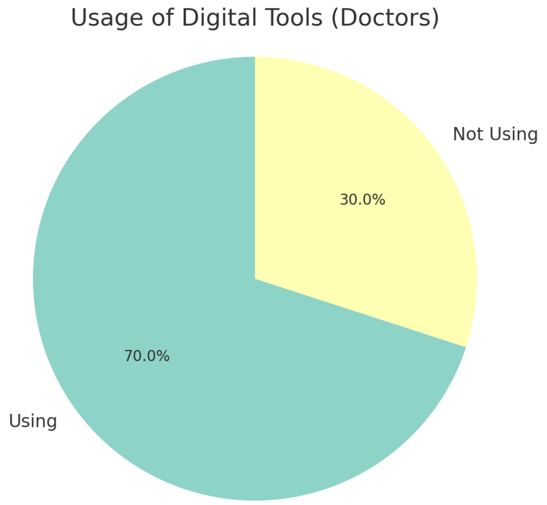

#### Support for Integration (Doctors)

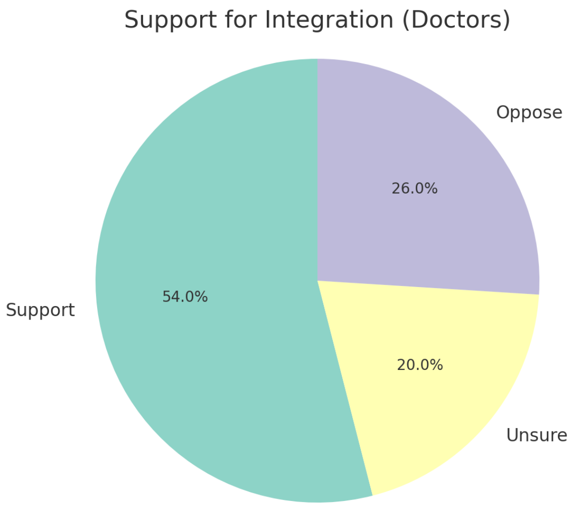


#### Challenges to Integration (Doctors)

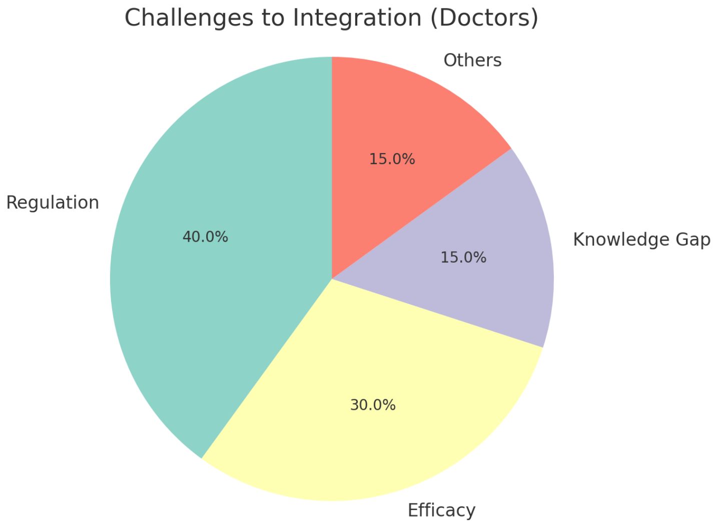

#### Perceived Benefits of Integration (Doctors)

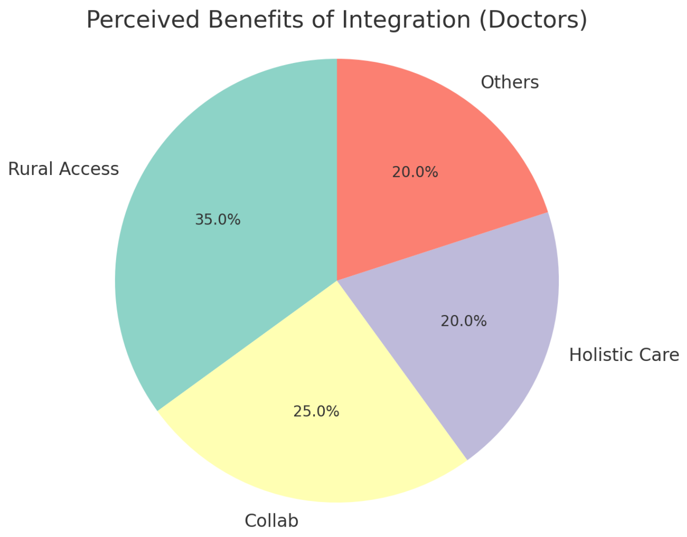
